# Estimating the early impact of vaccination against COVID-19 on deaths among elderly people in Brazil: analyses of routinely-collected data on vaccine coverage and mortality

**DOI:** 10.1101/2021.04.27.21256187

**Authors:** Cesar Victora, Marcia C Castro, Susie Gurzenda, Arnaldo Correia de Medeiros, Giovanny França, Aluisio J D Barros

## Abstract

**Background:** Vaccination against COVID-19 in Brazil started in January 2021, with health workers and the elderly as the priority groups. We assessed whether there was an impact of vaccinations on the mortality of elderly individuals in a context of wide transmission of the SARS-CoV-2 gamma (P.1) variant.

**Methods:** By May 27, 2021, 147238,414 COVID-19 deaths had been reported to the Brazilian Mortality Information System. Denominators for mortality rates were calculated by correcting population estimates for all-cause deaths reported in 2020. Proportionate mortality at ages 70-79 and 80+ years relative to deaths at all ages were calculated for deaths due to COVID-19 and to other causes, as were COVID-19 mortality rate ratios relative to individuals aged 0-69 years. Vaccine coverage data were obtained from the Ministry of Health. All results were tabulated by epidemiological weeks 1-19, 2021.

**Findings:** The proportion of all COVID-19 deaths at ages 80+ years was over 25% in weeks 1-6 and declined rapidly to 12.4% in week 19, whereas proportionate COVID-19 mortality for individuals aged 70-79 years started to decline by week 15. Trends in proportionate mortality due to other causes remained stable. Mortality rates were over 13 times higher in the 80+ years age group compared to that of 0-69 year olds up to week 6, and declined to 5.0 times in week 19. Vaccination coverage (first dose) of 90% was reached by week 9 for individuals aged 80+ years and by week 13 for those aged 70-79 years. Coronavac accounted for 65.4% and AstraZeneca for 29.8% of all doses administered in weeks 1-4, compared to 36.5% and 53.3% in weeks 15-19, respectively.

**Interpretation:** Rapid scaling up of vaccination coverage among elderly Brazilians was associated with important declines in relative mortality compared to younger individuals, in a setting where the gamma variant predominates. Had mortality rates among the elderly remained proportionate to what was observed up to week 6, an estimated additional 43,802 COVID-related deaths would have been expected up to week 19.

## Introduction

In early 2021, Brazil became the global epicenter of the COVID-19 pandemic ^1^ with an average of over 2,000 daily deaths in recent months.^2^ The gamma or P.1 variant, initially identified in Manaus in late 2020^3^ has rapidly spread throughout the country.^4^ Although genomic analyses are infrequent, in April and May 2021 the new variant accounted for three out of every four samples subjected to viral sequencing.^5^

Vaccination against COVID-19 was started in late January 2021, with two types of vaccines being offered: Coronavac (Sinovac, China) and AZD1222 (Oxford-AstraZeneca, UK). Vaccination has been initially targeted at four priority groups: health workers, the elderly (starting with those aged 85 years or more, and gradually vaccinating younger age groups), indigenous populations, and institutionalized individuals. By May 28, 41,478,005 Brazilians had received the first dose, and 19,604,603 the second dose. ^6^

Vaccination campaigns have been associated with reductions in hospital admissions and mortality among targeted population groups, in several of the early starting countries. ^7-9^ Yet, there is limited evidence on the efficacy of the two vaccines being delivered in Brazil against the gamma variant that currently accounts for the majority of cases in the country. Two observational studies among health care workers in Manaus^10^ and São Paulo^11^ suggested that the Coronavac provided partial protection against symptomatic illness in settings where gamma accounted for 75% and 47% of all infections, respectively, at the time of the study. Yet, there is growing concern that high SARS-CoV-2 incidence rates such as those observed in Brazil in early 2021 will lead to the appearance of new variants of concern as well as increase in the risk of vaccine escape. ^12^

To evaluate the real-life effectiveness of the vaccination campaign in Brazil, we analyzed time trends in mortality due to COVID-19 using a database of over 430,000 registered deaths. We hypothesized that mortality would fall more rapidly among the elderly, who were the initial target group of the vaccination campaign, than among younger Brazilians.

## Methods

Data on COVID-19 deaths were obtained from the Ministry of Health Mortality Information System^13^ including deaths reported until May 27, 2021. Coverage of the death registration system has been estimated at over 95% by 2010. ^14^ As of 2016, the Global Burden of Disease project assigned four out of five stars for the system’s coverage and quality of cause of death ascertainment,^15^ and by 2019 5.6% of all deaths were coded as due to ill-defined causes (França GA, unpublished data). We analyzed deaths for which the underlying cause was coded as B34.2, which included codes U07.1 (COVID-19, virus identified) and U07.2 (COVID-19, virus not identified).^16^ For 84% of 2021 deaths, presence of the virus was confirmed in a laboratory (preliminary results based on investigation of 163,637 deaths).

Data on COVID-19 vaccination coverage were obtained from a dataset made available by the Brazilian Ministry of Health.^6^ The data are updated daily and consist of an individual level dataset including personal information and information on the vaccination (type and dose) along with whether it is the first or second dose received and the priority group for the person vaccinated. The data used here were up to May 15, 2021.

Population estimates for July 1^st^ 2020 by single age and sex were obtained from the Brazilian Institute for Geography and Statistics (IBGE).^17^ Due to the excess mortality observed in 2020 and the higher COVID-19 mortality among the elderly,^18^ the population numbers from IBGE for 2020 are overestimated, particularly at older ages. Since vaccination started in Brazil in early January 2021, it is imperative to obtain an adjusted estimated population that more closely reflects the Brazilian population by the end of 2020. We considered the total deaths that were reported in 2020 (for all causes, as reported in the Mortality Information System), and the expected deaths as implied in the IBGE estimates. We excluded the additional number of deaths from the published 2020 estimates and used that adjusted population as the denominator in our analyses. All adjustments were made by age and sex. All calculations were done in R (R core team, 2020).

Mortality results were analyzed in two ways. First, we calculated proportionate mortality by dividing the number of COVID-19 deaths at ages 70-79 and 80+ years by the total number of COVID-19 deaths at all ages. Our main analyses described mortality by epidemiological week in 2021, which are supported by analyses by month of death during 2020. To investigate whether age-specific trends in proportionate mortality were specific to COVID-19 deaths, we also investigated trends due to other causes of death. Second, we calculated COVID-19 age-specific mortality rates by dividing the numbers of weekly deaths from the Mortality Information System by the estimated population by age group, as described above. Mortality rates at ages 70-79 and 80+ years were then divided by rates for the age range 0-9 years in the same week, resulting in mortality rate ratios.

Formal statistical tests were not performed as all results are based on the full country population, rather than samples. Analyses were carried out using Stata version 16 (StataCorp, College Station, TX, USA). All analyses were based on publicly available, anonymized databases.

## Results

From the beginning of the first epidemiological week in 2021 (January 3) to May 27, 238,414 deaths in the Mortality Information System were assigned to COVID-19 and 447,817 to other causes. Supplementary Table 1 shows the absolute number of COVID-19 deaths for epidemiological weeks 1-19 of 2021 (January 3 to May 15). There was rapid acceleration in deaths from week 9 (early March) when the gamma variant became the dominant strain. Results for weeks 17-19 (April 25 to May 15) are likely affected by registration delay but remain useful for comparing age-specific proportionate mortality and mortality rate ratios. Table 1 does not include deaths occurring after epidemiological week 19 (May 16 or later) as these are more markedly affected by delay than earlier deaths.

Figure 1 shows that proportionate COVID-19 mortality of individuals aged 80+ years fell rapidly from week 6 onwards, whereas proportionate mortality due to non-COVID causes remained relatively stable at just under 30%. Up to May 27, an additional 7,733 deaths had been reported for epidemiological weeks 20 and 21, of which 13.1% were among individuals aged 80+, a finding that is consistent with the levels achieved by week 15. Figure 1 also shows that proportionate mortality for individuals aged 70-79 years remained at around 25% up to week 15, when it started to decline sharply. For the same age group, proportionate mortality due to other causes remained stable at just over 20% of deaths at any age.

**Figure 1.**
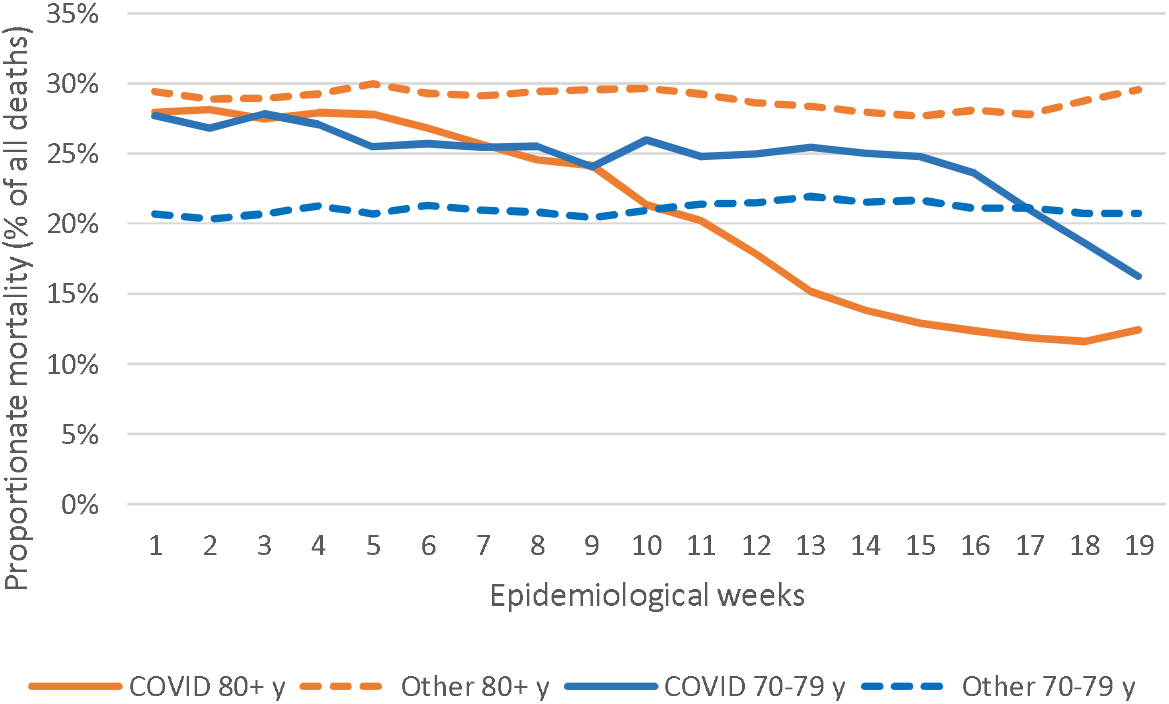
Proportionate mortality of individuals aged 70-79 and 80 or more years due to COVID-19 and all other causes, relative to deaths due to the same causes at all ages by epidemiological weeks, Brazil, 2021.

Supplementary Figure 1 shows that proportionate mortality at ages 80+ years fell in all regions of the country. The trend was less marked in the North region (where the Amazon is located) than in the rest of the country. Supplementary Figure 2 expands the time series by showing proportionate mortality based on 453,244 COVID-19 deaths that occurred since the beginning of the pandemic in the country. From May 2020 (when the monthly number of deaths exceeded 15,000) to January 2021, proportionate mortality at ages 80+ remained between 25% and 30%, with a sharp reduction starting in mid-February 2021. Proportionate mortality at ages 70-79 years remained above 20% until March 2021, with a substantial decline in April-May. Also showing data for 2020 and 2021, Supplementary Figure 3 demonstrates that the decline in proportionate mortality was observed for men and women, although proportionate mortality for women aged 80+ years tended to be higher than for men, likely due to higher life expectancy of women resulting in fewer deaths in those aged under 80 years.

Figure 2 shows time trends in mortality rate ratios using the age group 0-69 years as the reference. The mortality rate ratio for persons aged 80+ years fell from over 13.3 in January and early February to 8.0 in week 19. The decline in the rate ratio for ages 70-79 was more gradual, from 13.8 in week 1 to 5.0 in week 19. Mortality rate ratios for non-COVID causes remained stable over time.

**Figure 2.**
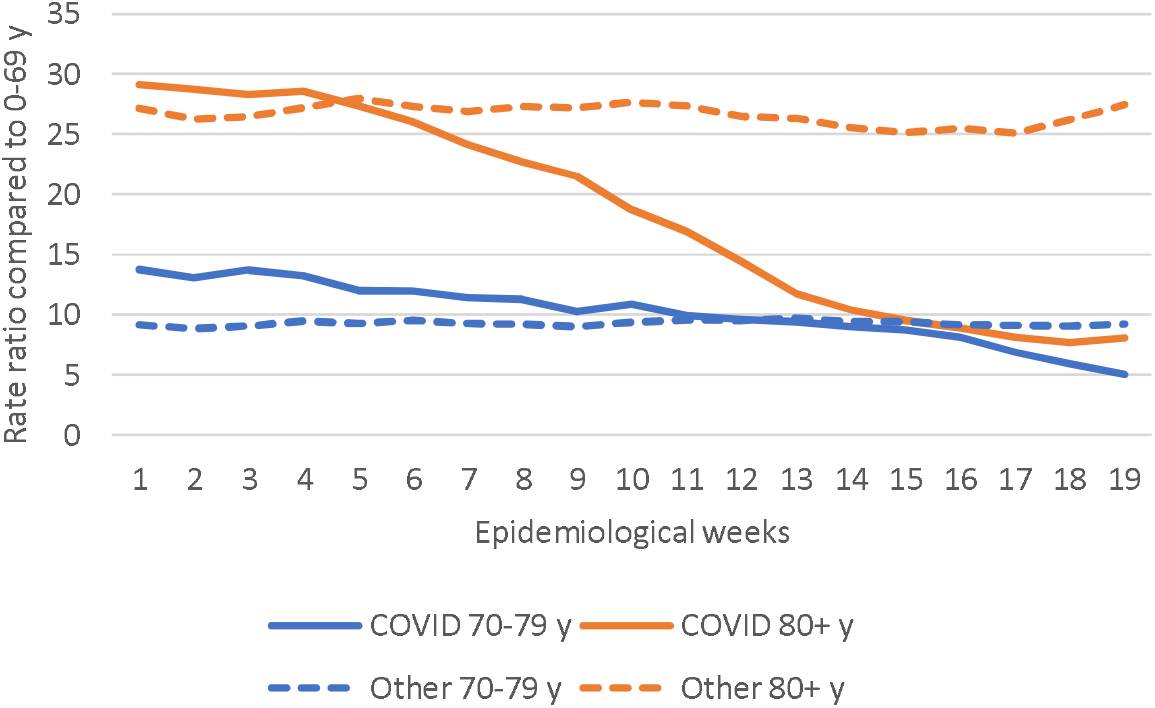
Mortality rate ratios: mortality rates at ages 70-79 and 80+ years divided by mortality rate at ages 0-69 years by epidemiological weeks, Brazil 2021.

Figure 3 shows vaccine coverage for individuals aged 70-79 and 80+ years over time. The increase in coverage was consistent with prioritization of older population groups, with 50% coverage reached for individuals aged 80+ years in the first half of February and over 80% by the second half, stabilizing at around 95% in March. For 70–79-year-olds, 50% coverage was reached by week 11 and 90% coverage by week 19. Coverage among younger age groups was largely restricted to priority groups including health workers, indigenous peoples and people living in institutions. In weeks 1-4, Coronavac accounted for 65.4% of all doses given and AstraZeneca for 29.8% whereas the corresponding percentages for weeks 15-19 were 36.5% and 53.3%. Pfizer/BioNTech (Germany) and Serum Institute (India) accounted for the remaining doses in the recent period.

**Figure 3.**
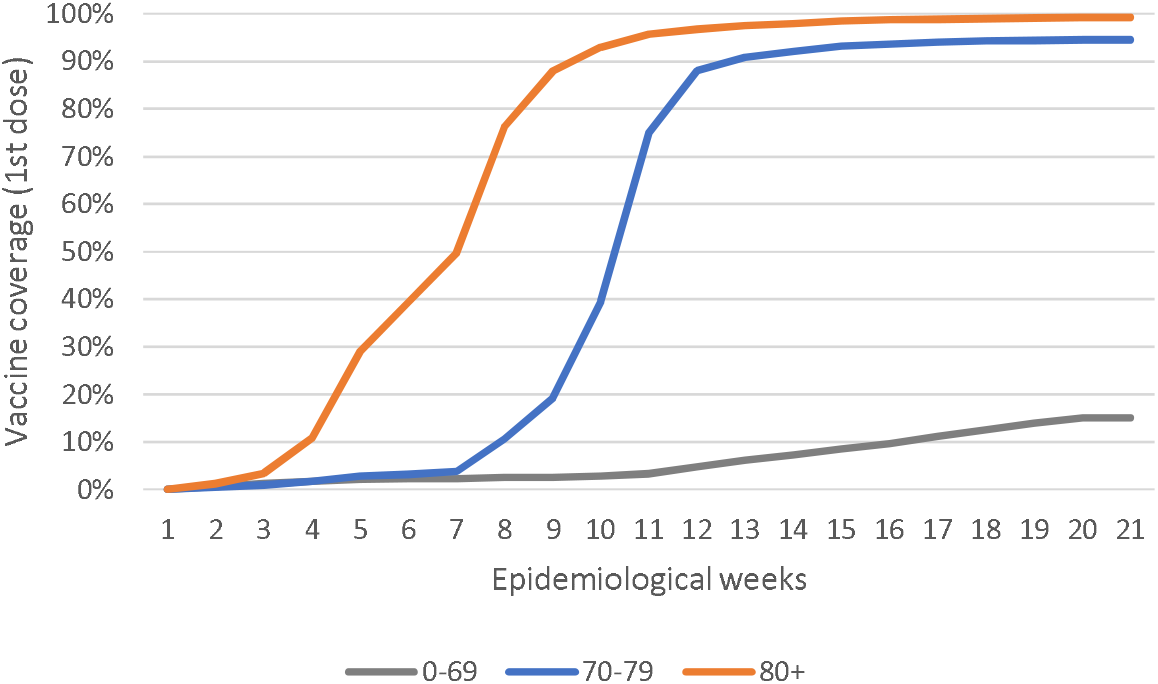
Covid-19 vaccination coverage (first dose) by age group by epidemiological week, Brazil, 2021.

The downturn in proportionate mortality due to COVID-19 started at about the sixth week of 2021. Had the number of deaths among individuals aged 80+ years continued to increase at the same rate as deaths among people aged 0-69 y, one would expect 70,015 such deaths during the 13-week period from mid-Feb to mid-May. Yet, 32,624 deaths were reported, or 37,401 fewer than expected under the scenario of similar trends as for the 0-69 years age group. A similar calculation was performed for deaths among 70–79-year-olds, among whom proportionate mortality started to decline around week 15. Compared to 13,838 deaths in weeks 15-19, 20,238 would be expected if mortality behaved similarly to that observed for 0– 69-year-olds. Adding the two estimates, 43,802 deaths may have been avoided by the decline in mortality among the elderly.

## Discussion

We found evidence that, although dissemination of the gamma variant led to increases in reported COVID-19 death at all ages, the proportion of deaths among the elderly started to fall rapidly from the second half of February 2021. This proportion had been stable at around 25%-30% since the beginning of the epidemic in early 2020 but is now below 13% in May 2021.

Estimates of proportionate mortality must be interpreted with caution. We now describe how we handled potential caveats in these analyses.

First, the absolute number of deaths in the elderly may be reduced due to smaller number of persons at risk, resulting from high mortality in 2020 due to COVID-19 and other causes. In an estimated population of approximately 815 thousand Brazilians aged 90+ years in 2020, there were approximately 144 thousand deaths in the calendar year, of which about 10% were reported as being caused by COVID-19. To address this potential caveat, our calculations of mortality rates for 2021 were based on population estimates at the beginning of the year from which all-cause deaths had already been deducted.

Second, proportionate mortality may be spuriously reduced among the elderly if the gamma variant of concern disproportionally affected younger individuals, either in terms of infection rates or of infection-fatality rates. The EPICOVID-19 study has been monitoring prevalence of antibodies against SARS-CoV-2 through household surveys in nine large cities in the state of Rio Grande do Sul since April 2020. In early February 2021, antibody prevalence levels were 9.6%, 11.3%, 10.0% and 8.3% for unvaccinated individuals aged 10-19, 20-39, 40-59, and 60+ years, respectively (AJD Barros, personal communication). The state has been strongly affected by the recent pandemic wave, yet there is no evidence of important age patterns in antibody prevalence.

Thirdly, our results based on ratios of mortality rates closely mirror the findings from the proportionate mortality analyses, showing that the rate ratio for individuals aged 80+ relative to those aged 0-69 years fell from 13.3 in January to 8.0 in April.

Lastly, our analyses of deaths due to causes other than COVID-19 showed that proportionate mortality and mortality rate ratios for the elderly remained stable over time, thus supporting the specificity of an impact on COVID-19 deaths.

Another potential limitation of our analyses is the underreporting of deaths and delays in reporting. Delays are particularly relevant for estimating mortality rates for recent periods, as only deaths that reached the system by May 27 were included. However, proportionate mortality by age groups would only be affected if delays varied systematically with age, which is unlikely. As discussed in the Introduction, the overall coverage of mortality statistics has been very high in Brazil for many years, and ill-defined causes represent 5.6% of all deaths. The mortality database for the present analyses includes approximately 30% more deaths than the SIVEP-Gripe database on hospital admissions and mortality that has been employed in previous analyses of COVID-19 deaths in Brazil.^18-20^

However, there is evidence that the excess mortality during 2020 relative to earlier years was not fully explained by deaths due to COVID-19. It is likely that some of such deaths were reported as having been due to other causes or to ill-defined conditions, but it is also possible that increases in non-COVID-19 deaths were because health services were under stress due to the large COVID-19 case load. Unless reporting patterns varied by age or calendar time, this limitation is unlikely to affect the present results particularly in light of the present finding that age patterns in deaths assigned to non-COVID causes remained stable.

The decline in mortality was observed for both sexes. Proportionate mortality at older ages was higher among women than for men, which is compatible with higher case-fatality of younger male adults, possibly related to comorbidities, given that existing serological surveys do not suggest differences in infection prevalence by sex.^21,22^ The reductions in proportionate mortality were very similar across four of the five regions of the country. A decline was also observed in the fifth region (Northern Brazil including the Amazon), but proportionate mortality was lower at the beginning of the year than in the rest of the country, and the decline started later than in the rest of the country. The North region has been badly hit by the first and second waves of the pandemic, and high prevalence, high case-fatality, and the limited availability of health services in this region^23^ may have led to a larger number of deaths among young adults. Even before the pandemic, life expectancy at birth in the North region was the shortest in the country at 72.9 years, compared to 73.9, 78.3, 78.6 and 75.8 in the Northeast, Southeast, South and Center-West, respectively.^17^

The most likely explanation for the observed reductions in proportionate mortality and in rate ratios for the elderly is the rapid increase in vaccination coverage in these age groups, as has been described for other parts of the world. ^7-9^ The increase in vaccine coverage preceded the decline in mortality, and the decline at ages 80+ years preceded the decline at ages 70-79 years, which is in accordance with the vaccination calendar.

Our results are original in the sense that none of existing population-based mortality studies were carried out in a setting where the gamma variant is predominant. Recent observational studies in vaccinated health workers in Manaus and São Paulo^10,11^ had already suggested that Coronavac provided some degree of protection against symptomatic illness in settings where gamma was prevalent. Coronavac accounted for most vaccinations in the 80+ years age group, who were immunized in January and February, with AstraZeneca vaccine accounting for the majority of recent doses. Individuals who received the latter are so far protected by a single dose given that the second dose is provided 12 weeks after the first, whereas the second dose of Coronavac has already been administered to a very high proportion of individuals aged 80+ years ^24^ as doses are given four weeks apart. The health worker study in São Paulo suggested that the number of cases started to drop after the first Coronavac dose, which is compatible with our findings. ^11^ This is supported by the results of a recent mass vaccination trial with Coronavac in the town of Serrana (population 27,000) carried out by Instituto Butantan. Following high coverage with Coronavac in early 2021, reductions of 86% in admissions and 95% in deaths were observed in the town by the end of May.^25^

We attempted to provide an approximate estimate of lives saved among elderly Brazilians in the eight-week period since vaccination was accelerated throughout the country. The figure of over 40 thousand deaths averted is likely an underestimate, because it does not take into account lives saved among other priority groups for vaccination, such as health workers and indigenous populations. Also, by using the mortality in ages 0-69 years to predict expected deaths among those aged 70+ years, we are not accounting for lives saved by the vaccine among younger age groups – e.g., 60-69-year-olds - for whom coverage also increased, albeit at a slower rate. Although it is not possible to make strong causal arguments on the basis of the data available for our analyses, our findings are consistent with the results of efficacy trials for both vaccines, and with observational studies in high-risk groups of health workers.^10,11^

The main contribution brought by the present analyses is to provide large-scale supporting evidence for effectiveness of vaccination in a setting with wide circulation of the gamma variant. Because compliance with non-pharmaceutical interventions such as social distancing and mask use is limited in most of the country, rapid scaling up of vaccination remains as the most promising approach for controlling the pandemic in a country where almost 500,000 lives have already been lost to COVID-19.

## Data Availability

The analyses are based on publicly available data at the www.saude.gov.br website (detailed information on the exact urls is provided in the reference list).

## Supplementary materials

### Table and Figure Captions

**Supplementary figure 1.**
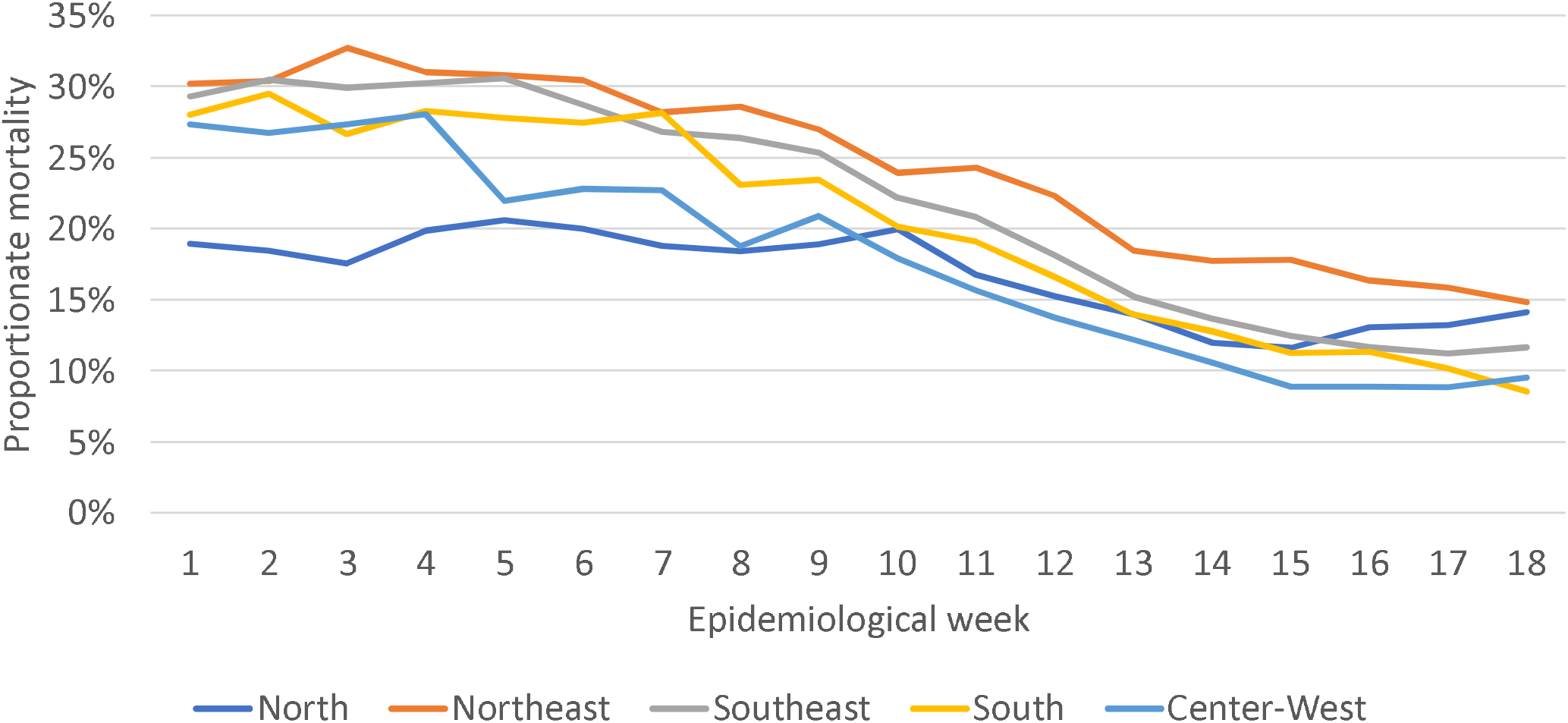
Proportionate mortality of individuals aged 80+ years due to COVID-19 relative to deaths at all ages due to COVID-19 by region and epidemiological week. Brazil, January to April 2021.

**Supplementary figure 2.**
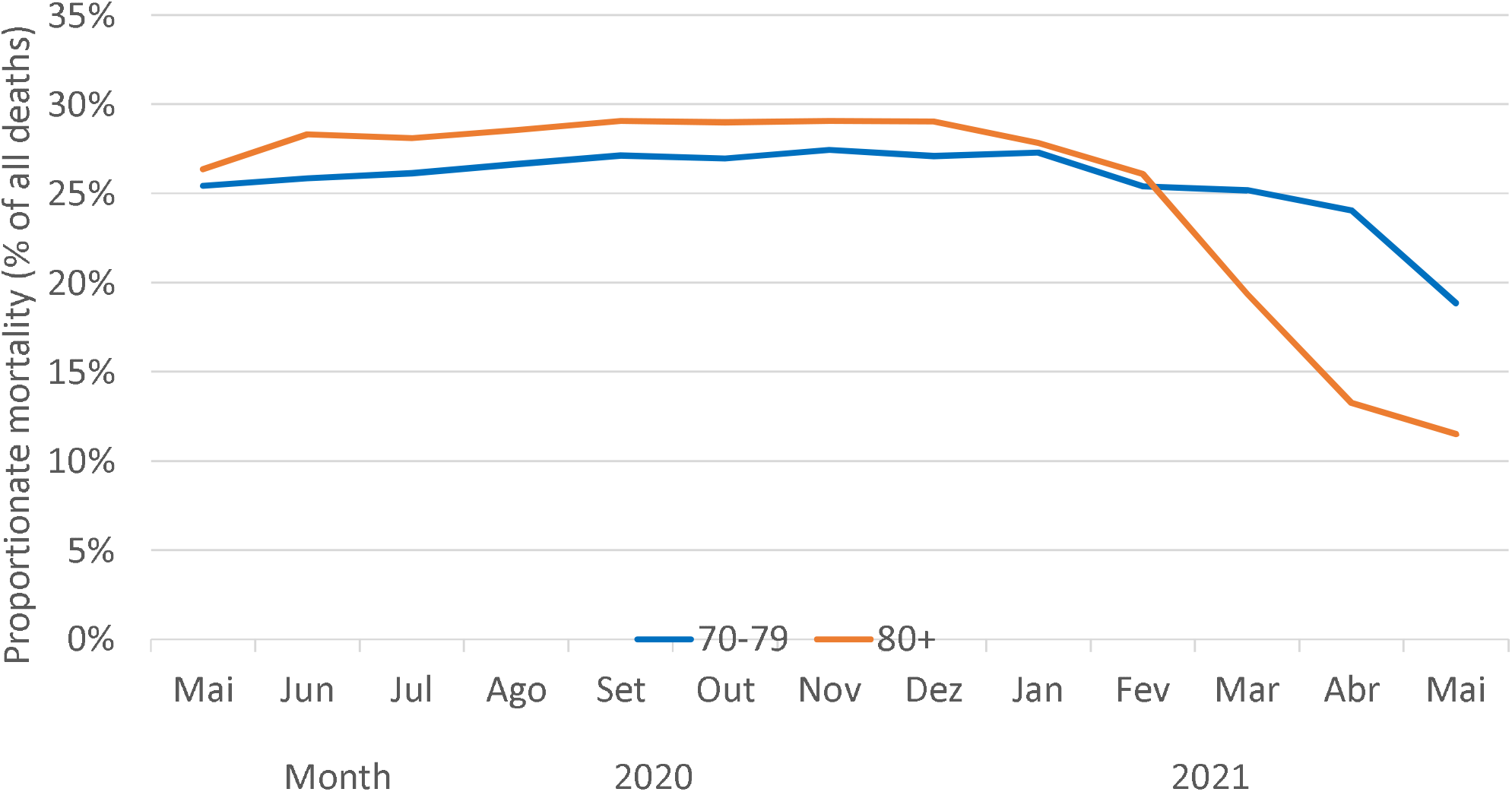
Proportionate mortality due to COVID-19 of individuals aged 70-79 and 80+ years relative to deaths at all ages due to COVID-19 by month. Brazil, May 2020 to May 2021.

**Supplementary figure 3.**
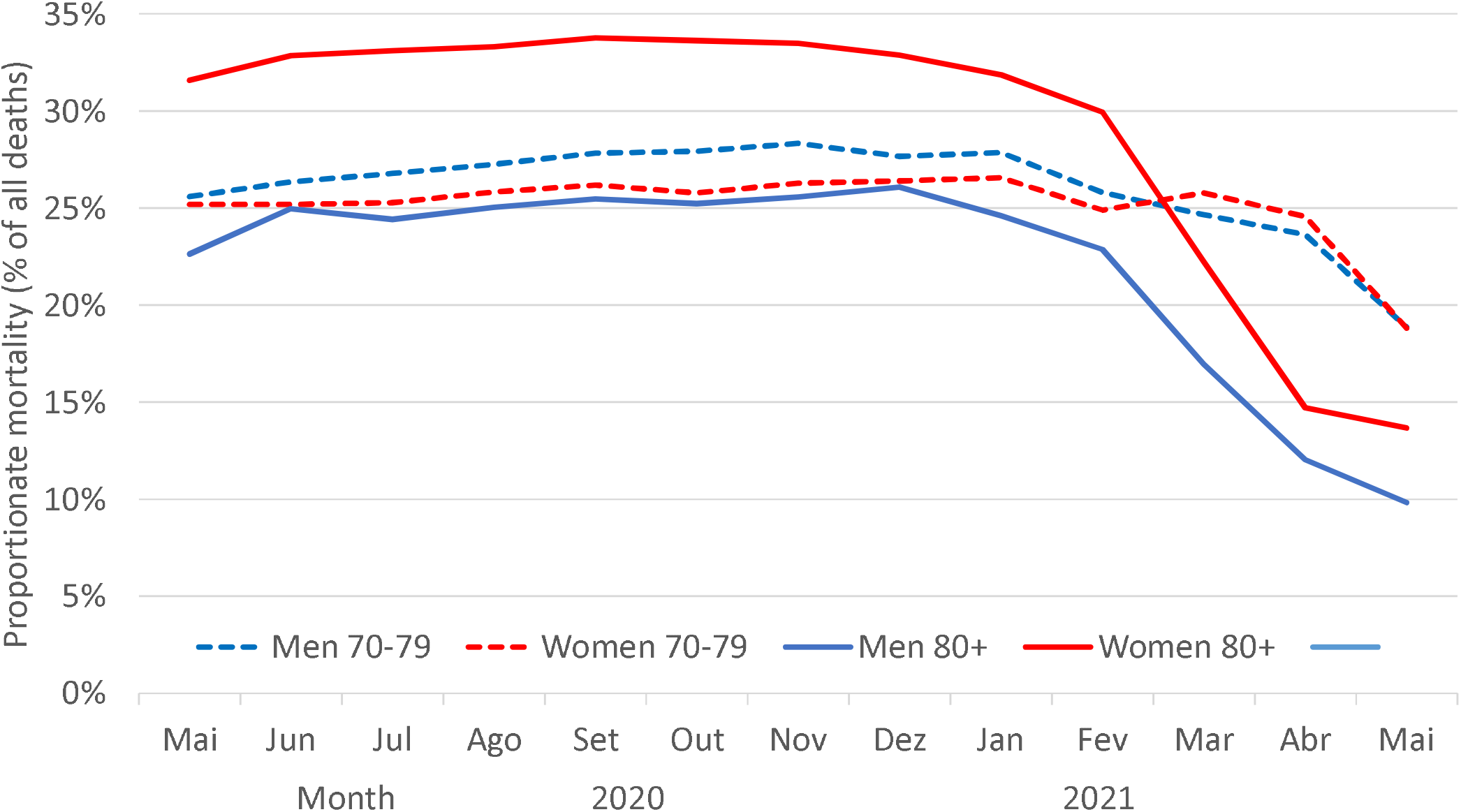
Sex-specific proportionate mortality due to COVID-19 of individuals aged 70-79 and 80+ years relative to deaths at all ages due to COVID-19 by month. Brazil, May 2020 to May 2021.

**Supplementary table 1.**
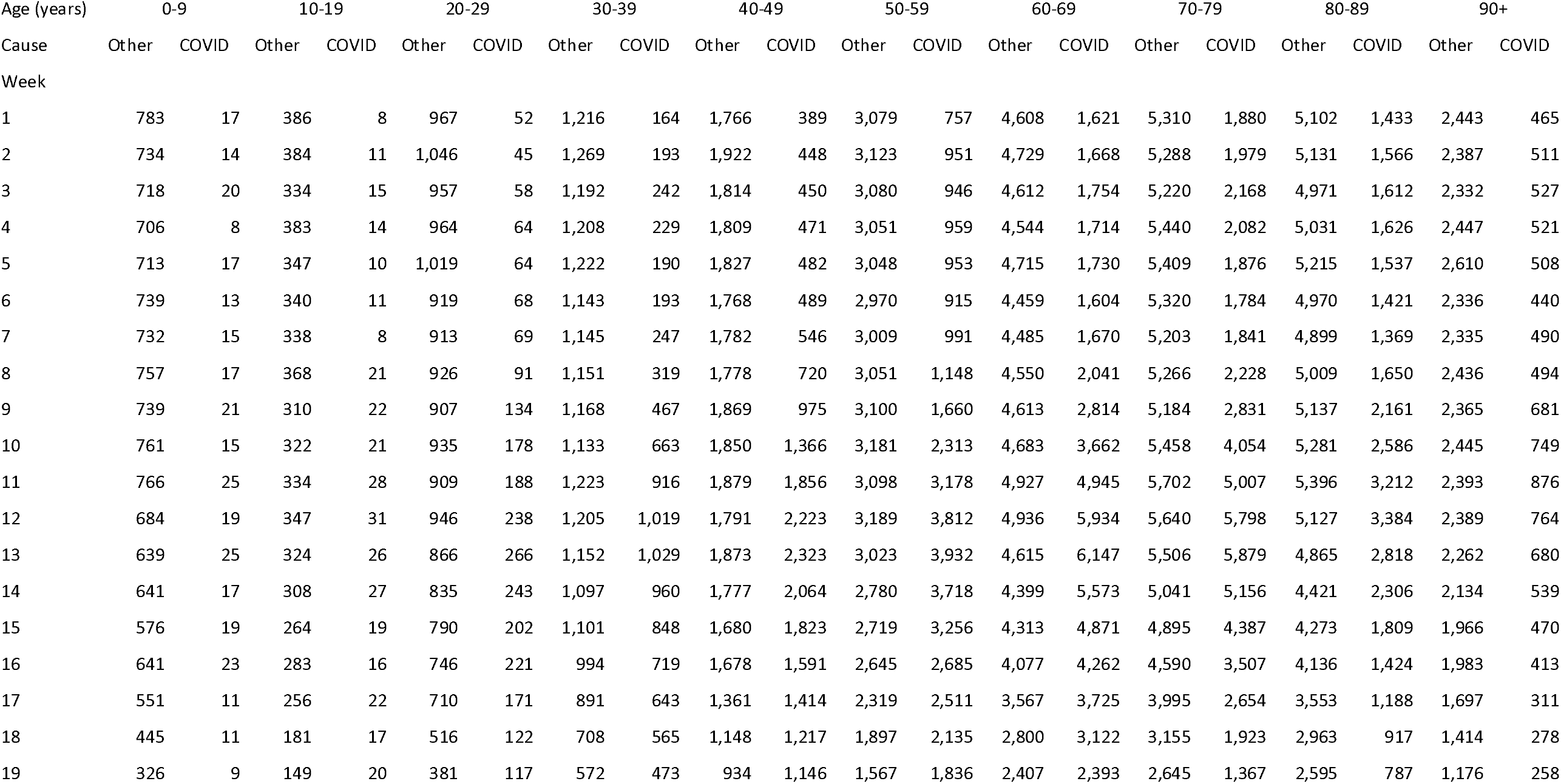
Absolute number of deaths due to COVID-19 and to all other causes by epidemiological week according to age groups. Brazil,2021.

| Age (years) | 0-9 |  | 10-19 |  | 20-29 |  | 30-39 |  | 40-49 |  | 50-59 |  | 60-69 |  | 70-79 |  | 80-89 |  | 90+ |  |
| --- | --- | --- | --- | --- | --- | --- | --- | --- | --- | --- | --- | --- | --- | --- | --- | --- | --- | --- | --- | --- |
| Cause | Other | COVID | Other | COVID | Other | COVID | Other | COVID | Other | COVID | Other | COVID | Other | COVID | Other | COVID | Other | COVID | Other | COVID |
| Week |  |  |  |  |  |  |  |  |  |  |  |  |  |  |  |  |  |  |  |  |
| 1 | 783 | 17 | 386 | 8 | 967 | 52 | 1,216 | 164 | 1,766 | 389 | 3,079 | 757 | 4,608 | 1,621 | 5,310 | 1,880 | 5,102 | 1,433 | 2,443 | 465 |
| 2 | 734 | 14 | 384 | 11 | 1,046 | 45 | 1,269 | 193 | 1,922 | 448 | 3,123 | 951 | 4,729 | 1,668 | 5,288 | 1,979 | 5,131 | 1,566 | 2,387 | 511 |
| 3 | 718 | 20 | 334 | 15 | 957 | 58 | 1,192 | 242 | 1,814 | 450 | 3,080 | 946 | 4,612 | 1,754 | 5,220 | 2,168 | 4,971 | 1,612 | 2,332 | 527 |
| 4 | 706 | 8 | 383 | 14 | 964 | 64 | 1,208 | 229 | 1,809 | 471 | 3,051 | 959 | 4,544 | 1,714 | 5,440 | 2,082 | 5,031 | 1,626 | 2,447 | 521 |
| 5 | 713 | 17 | 347 | 10 | 1,019 | 64 | 1,222 | 190 | 1,827 | 482 | 3,048 | 953 | 4,715 | 1,730 | 5,409 | 1,876 | 5,215 | 1,537 | 2,610 | 508 |
| 6 | 739 | 13 | 340 | 11 | 919 | 68 | 1,143 | 193 | 1,768 | 489 | 2,970 | 915 | 4,459 | 1,604 | 5,320 | 1,784 | 4,970 | 1,421 | 2,336 | 440 |
| 7 | 732 | 15 | 338 | 8 | 913 | 69 | 1,145 | 247 | 1,782 | 546 | 3,009 | 991 | 4,485 | 1,670 | 5,203 | 1,841 | 4,899 | 1,369 | 2,335 | 490 |
| 8 | 757 | 17 | 368 | 21 | 926 | 91 | 1,151 | 319 | 1,778 | 720 | 3,051 | 1,148 | 4,550 | 2,041 | 5,266 | 2,228 | 5,009 | 1,650 | 2,436 | 494 |
| 9 | 739 | 21 | 310 | 22 | 907 | 134 | 1,168 | 467 | 1,869 | 975 | 3,100 | 1,660 | 4,613 | 2,814 | 5,184 | 2,831 | 5,137 | 2,161 | 2,365 | 681 |
| 10 | 761 | 15 | 322 | 21 | 935 | 178 | 1,133 | 663 | 1,850 | 1,366 | 3,181 | 2,313 | 4,683 | 3,662 | 5,458 | 4,054 | 5,281 | 2,586 | 2,445 | 749 |
| 11 | 766 | 25 | 334 | 28 | 909 | 188 | 1,223 | 916 | 1,879 | 1,856 | 3,098 | 3,178 | 4,927 | 4,945 | 5,702 | 5,007 | 5,396 | 3,212 | 2,393 | 876 |
| 12 | 684 | 19 | 347 | 31 | 946 | 238 | 1,205 | 1,019 | 1,791 | 2,223 | 3,189 | 3,812 | 4,936 | 5,934 | 5,640 | 5,798 | 5,127 | 3,384 | 2,389 | 764 |
| 13 | 639 | 25 | 324 | 26 | 866 | 266 | 1,152 | 1,029 | 1,873 | 2,323 | 3,023 | 3,932 | 4,615 | 6,147 | 5,506 | 5,879 | 4,865 | 2,818 | 2,262 | 680 |
| 14 | 641 | 17 | 308 | 27 | 835 | 243 | 1,097 | 960 | 1,777 | 2,064 | 2,780 | 3,718 | 4,399 | 5,573 | 5,041 | 5,156 | 4,421 | 2,306 | 2,134 | 539 |
| 15 | 576 | 19 | 264 | 19 | 790 | 202 | 1,101 | 848 | 1,680 | 1,823 | 2,719 | 3,256 | 4,313 | 4,871 | 4,895 | 4,387 | 4,273 | 1,809 | 1,966 | 470 |
| 16 | 641 | 23 | 283 | 16 | 746 | 221 | 994 | 719 | 1,678 | 1,591 | 2,645 | 2,685 | 4,077 | 4,262 | 4,590 | 3,507 | 4,136 | 1,424 | 1,983 | 413 |
| 17 | 551 | 11 | 256 | 22 | 710 | 171 | 891 | 643 | 1,361 | 1,414 | 2,319 | 2,511 | 3,567 | 3,725 | 3,995 | 2,654 | 3,553 | 1,188 | 1,697 | 311 |
| 18 | 445 | 11 | 181 | 17 | 516 | 122 | 708 | 565 | 1,148 | 1,217 | 1,897 | 2,135 | 2,800 | 3,122 | 3,155 | 1,923 | 2,963 | 917 | 1,414 | 278 |
| 19 | 326 | 9 | 149 | 20 | 381 | 117 | 572 | 473 | 934 | 1,146 | 1,567 | 1,836 | 2,407 | 2,393 | 2,645 | 1,367 | 2,595 | 787 | 1,176 | 258 |

## References

1. Castro MC, Kim S, Barberia L, et al. Spatiotemporal pattern of COVID-19 spread in Brazil. Science 2021.

2. Brasil. Ministério da Saúde. COVID-19 Painel Coronavírus. https://covid.saude.gov.br/ (accessed June 16, 2021).

3. Faria NR, Mellan TA, Whittaker C, et al. Genomics and epidemiology of the P.1 SARS-CoV-2 lineage in Manaus, Brazil. Science 2021).

4. Fundação Oswaldo Cruz. Fiocruz detecta mutação associada a variantes de preocupação no país. March 4, 2021 2021. https://portal.fiocruz.br/noticia/fiocruz-detecta-mutacao-associada-variantes-de-preocupacao-no-pais (accessed June 15, 2021).

5. GISAID. Genomic epidemiology of hCoV-19. 2021). www.gisaid.org (accessed June 15, 2021).

6. Brasil. Ministério da Saúde. Opendatasus. Campanha Nacional de Vacinação contra Covid-19. https://opendatasus.saude.gov.br/dataset/covid-19-vacinacao (accessed June 15, 2021).

7. Vasileiou E, Simpson CR, Robertson C, et al. Effectiveness of First Dose of COVID-19 Vaccines Against Hospital Admissions in Scotland: National Prospective Cohort Study of 5.4 Million People. SSRN 2021).

8. Israel Ministry of Health. Effectiveness Data of the COVID-19 Vaccine Collected in Israel until 13.2.2021. https://www.gov.il/en/departments/news/20022021-01 (accessed June 15, 2021).

9. Bernal JL, Andrews N, Gower C, et al. Early effectiveness of COVID-19 vaccination with BNT162b2 mRNA vaccine and ChAdOx1 adenovirus vector vaccine on symptomatic disease, hospitalisations and mortality in older adults in England. medRxiv: 2021.03.01.21252652.

10. Hitchings MDT, Ranzani OT, Scaramuzzini Torres MS, et al. Effectiveness of CoronaVac in the setting of high SARS-CoV-2 P.1 variant transmission in Brazil: A test-negative case-control study. MedRxiv: 2021.04.07.21255081.

11. de Faria E, Guedes AR, Oliveira MS, et al. Performance of vaccination with CoronaVac in a cohort of healthcare workers (HCW) - preliminary report. medRxiv: 2021.04.12.21255308.

12. Thompson RN, Hill EM, Gog JR. SARS-CoV-2 incidence and vaccine escape. The Lancet Infectious diseases 2021: S1473-3099(21)00202-4.

13. Brasil. Ministério da Saúde. SIM. Sistema de Informações de Mortalidade. http://www2.datasus.gov.br/DATASUS/index.php?area=060701 (accessed June 15, 2021).

14. Lima EECd, Queiroz BL. Evolution of the deaths registry system in Brazil: associations with changes in the mortality profile, under-registration of death counts, and ill-defined causes of death. Cadernos de Saúde Pública 2014; 30: 1721-30.

15. Marinho F, de Azeredo Passos VM, Carvalho Malta D, et al. Burden of disease in Brazil, 1990–2016: a systematic subnational analysis for the Global Burden of Disease Study 2016. The Lancet 2018; 392(10149): 760–75.

16. World Health Organization. Emergency use ICD codes for COVID-19 disease outbreak. https://www.who.int/standards/classifications/classification-of-diseases/emergency-use-icd-codes-for-covid-19-disease-outbreak (accessed June 15, 2021).

17. Instituto Brasileiro de Geografia e Estatística. Projeções da População. https://www.ibge.gov.br/estatisticas/sociais/populacao/9109-projecao-da-populacao.html?=&t=o-que-e (accessed June 15, 2021).

18. Baqui P, Bica I, Marra V, Ercole A, van der Schaar M. Ethnic and regional variations in hospital mortality from COVID-19 in Brazil: a cross-sectional observational study. Lancet Glob Health 2020; 8(8): e1018–e26.

19. Niquini RP, Lana RM, Pacheco AG, et al. Description and comparison of demographic characteristics and comorbidities in SARI from COVID-19, SARI from influenza, and the Brazilian general population. Cad Saude Publica 2020; 36(7): e00149420.

20. Ranzani OT, Bastos LSL, Gelli JGM, et al. Characterisation of the first 250,000 hospital admissions for COVID-19 in Brazil: a retrospective analysis of nationwide data. Lancet Respir Med 2021; 9(4): 407–18.

21. Hallal PC, Hartwig FP, Horta BL, et al. SARS-CoV-2 antibody prevalence in Brazil: results from two successive nationwide serological household surveys. Lancet Glob Health 2020; 8(11): e1390–e8.

22. Hallal PC, et al. Slow spread of SARS-CoV-2 in Southern Brazil over a six-month period: Report on eight sequential statewide serological surveys including 35,611 participants. Am J Public Health 2021; (in press).

23. Albuquerque MVd, Viana ALdÁ, Lima LDd, Ferreira MP, Fusaro ER, Iozzi FL. Desigualdades regionais na saúde: mudanças observadas no Brasil de 2000 a 2016. Ciência & Saúde Coletiva 2017; 22: 1055-64.

24. Brasil. Ministério da Saúde. COVID-19 Vacinação - Distribuição de Vacinas. https://qsprod.saude.gov.br/extensions/DEMAS_C19VAC_Distr/DEMAS_C19VAC_Distr.html (accessed June 15, 2021).

25. Instituto Butantan. Projeto S: imunização em Serrana faz casos de Covid-19 despencarem 80% e mortes, 95%. https://butantan.gov.br/noticias/projeto-s-imunizacao-em-serrana-faz-casos-de-covid-19-despencarem-80-e-mortes-95 (accessed June 15, 2021).

